# Towards predicting PTSD symptom severity using portable EEG-derived biomarkers

**DOI:** 10.1101/2024.07.17.24310570

**Authors:** Ashritha Peddi, Mohammad S. E. Sendi, Sean T. Minton, Cecilia A. Hinojosa, Emma West, Ryan Langhinrichsen-Rohling, Kerry J. Ressler, Vince D. Calhoun, Sanne J.H. van Rooij

## Abstract

Posttraumatic Stress Disorder (PTSD) is a heterogeneous mental health disorder that occurs following traumatic experience. Understanding its neurobiological basis is crucial to advance early diagnosis and treatment. Electroencephalography (EEG) can be used to explore the neurobiological basis of PTSD. However, only limited research has explored mobile EEG, which is important for scalability. This proof-of-concept study delves into mobile EEG-derived biomarkers for PTSD and their potential implications. Over four weeks, we measured PTSD symptoms using the PTSD checklist for DSM-5 (PCL-5) at multiple timepoints, and we recorded multiple EEG sessions from 21 individuals using a mobile EEG device. In total, we captured 38 EEG sessions, each comprising two recordings that lasted approximately 180 seconds, to evaluate reproducibility. Next, we extracted Shannon entropy, as a measure of the randomness or unpredictability of the signal and spectral power for the fronto-temporal regions of interest, including electrodes at AF3, AF4, T7, and T8 for each EEG recording session. We calculated the partial correlation between the EEG variables and PCL-5 measured closest to the EEG session, using age, sex, and the grouping variable ‘batch’ as covariates. We observed a significant negative correlation between Shannon entropy in fronto-temporal regions and PCL-5 scores. Specifically, this association was evident in the AF3 (*r* = -0.456, FDR-corrected *p* = 0.01), AF4 (*r* = -0.362, FDR-corrected *p* = 0.04), and T7 (*r* = -0.472, FDR-corrected *p* = 0.01) regions. Additionally, we found a significant negative association between the alpha power estimated from AF4 and PCL-5 (*r*=-0.429, FDR-corrected *p*=0.04). Our findings suggest that EEG data acquired using a mobile EEG device is associated with PTSD symptom severity, offering valuable insights into the neurobiological mechanisms underlying PTSD.

## 1. Introduction

Posttraumatic stress disorder (PTSD) is a complex mental health condition that may develop in response to exposure to traumatic events. Approximately 6% of people in the United States will suffer from PTSD at some time in their life. This disorder is accompanied by significant distress, impairment in everyday functioning, and social and emotional well-being issues ^1^. Its diagnosis and treatment present significant challenges due to the diversity of symptoms and their subjective nature ^2^. To address these issues, it is critical to develop improved knowledge of the neurobiological basis of PTSD and to investigate novel quantitative biomarkers to diagnosis and measure response to treatment.

Electroencephalography (EEG) is a non-invasive method that measures electrical activity in the brain through sensors placed on the scalp. EEG has been developed as a useful tool for investigating the neural signatures of a variety of neurological and psychiatric diseases, including PTSD ^3^. For example, multiple recent studies showed that functional connectivity in the theta band can predict more severe PTSD symptoms ^4,5^. Moreover, analyses of EEG data from patients with PTSD indicate abnormalities such as a widespread reduction in alpha wave power ^6^. This reduction in alpha power may reflect decreased cortical inhibition or increased cortical excitability, which are common in PTSD and may contribute to symptoms such as hyperarousal and impaired cognitive processing. Additionally, a recent study using time-dependent Hurst analysis on EEG signals revealed that healthy controls (HC) exhibited greater complexity in their brain activity patterns, as evidenced by the Hurst exponent in the F3 channel (frontal region). This significantly differentiated them from individuals with combat-related PTSD, highlighting the diagnostic potential of the Hurst exponent estimated from EEG^7^. It suggests that individuals with PTSD exhibit more predictable and less variable brain activity patterns.

EEG studies have traditionally relied on conventional EEG instruments, which demand an extensive setup including skin preparation, electrode positioning, gel application, montage selection, and device connection ^8^. These standard procedures are not only expensive and time-consuming but can also be uncomfortable for patients, often making them feel out of their natural surroundings. Given the profound clinical insights derived from EEG data and the inherent limitations of these traditional setups, there is a burgeoning interest in mobile EEG systems ^9^. These systems offer the allure of prolonged monitoring in a user-centric design without compromising the data quality compared to their conventional counterparts ^10^. The development of mobile EEG devices represents a significant step forward in brain research and clinical practices. Their portability ensures they can be used in diverse settings, breaking free from the limitations of standard clinical environments, and expanding scalability ^11^. Such adaptability is crucial for broader application in natural settings, where real-time data gathering in these settings can offer invaluable insights into the disorder’s neural foundations ^12^. Furthermore, the ability of these devices to monitor neural signatures continuously or at frequent intervals can be instrumental in the early detection of symptom changes. This early identification is crucial for timely interventions, potentially preventing severe disease progression or symptom aggravation.

While mobile EEG offers an intriguing approach to addressing diverse neuroscience questions, the potential of mobile EEG technology to predict the severity of symptoms in neuropsychiatric disorders like PTSD remains a prospective area for future research. The main objectives of our current study are to 1) monitor PTSD symptom severity using mobile EEG technology, and 2) determine if EEG entropy is associated with PTSD severity. We hypothesize that the use of mobile EEG devices will provide valuable insights into the neurobiological aspects of the disorder, offering a practical and effective alternative for understanding and monitoring PTSD symptoms. Additionally, we hypothesize that there will be a significant association between entropy measures derived from EEG signals and the severity of PTSD symptoms. Specifically, we expect that higher PTSD severity will correspond to lower entropy values, indicating a more predictable and less complex brain activity pattern.

In this study, we specifically examined Shannon entropy in four EEG channels: AF3, T7, T8, and AF4. These channels are predominantly located in the fronto-temporal region of the brain. The fronto-temporal lobe plays a crucial role in emotional processing, memory, and cognitive functions, which are often compromised in individuals with PTSD^13^. Also, entropy has proven particularly effective in analyzing EEG signals to distinguish various emotions based on their inherent irregularities^14^. For example, a previous study introduced an emotion recognition system utilizing EEG channels FP1, FP2, T3, T4, and Pz. Using approximate and wavelet entropies extracted from these channels, emotional states were effectively classified with a 73.25% accuracy via support vector machine, highlighting a reduced EEG complexity during emotional experiences^14^. Another study employed Shannon entropy to analyze EEG patterns and discerned those acute stressors, such as foot shock stimulation, instigated discernible shifts in EEG entropy during both REM and NREM sleep phases in rats. This finding underscores the potential of Shannon entropy in identifying stress-related sleep disturbances^15^.

We also explored the spectral power in the alpha band (9-13 Hz) for the following reasons: 1) Previous research has demonstrated that among delta, theta, alpha, and beta bands, the alpha and beta power measurements collected using dry/mobile EEG headsets are comparable to those obtained with conventional EEG devices^16,17^. Also, a study focusing on the Emotiv device at the AF3 and AF4 locations indicated that, among the theta, alpha, and beta bands, the alpha band signal showed the greatest comparability with conventional devices ^18^. 2) Given our sampling frequency of 128 Hz, we believe our power estimation would be more accurate in the alpha band comparing with beta. Particularly in the higher range of the beta band, our power estimates are likely to be less reliable. 3) Previous studies have highlighted the significant role of alpha power in relation to PTSD^19,20^. Additionally, the alpha band has been associated with cognitive processes such as attention, relaxation, and mental states, making it a relevant target for investigation in the context of PTSD. Overall, we concentrated on extracting the spectral power and Shannon entropy in the alpha band from fronto-temporal regions, specifically the AF3, AF4, T7, and T8 electrodes.

## 2. Method

### 2.1 Participants

In this study, participants (N=21, age: *M*=43.9, *SD*=13.9) were drawn from a broader double-blind clinical trial investigating the effect of transcranial magnetic stimulation (TMS) on PTSD symptoms and biomarkers (https://clinicaltrials.gov/study/NCT04563078). In the TMS study, individuals underwent either active or sham 1Hz TMS to the right dorsolateral prefrontal cortex for 20 sessions (2 sessions per day) over a two-week period. The participant group comprised four males and seventeen females. Demographic and clinical characteristics of the samples are summarized in Table 1. Inclusion criteria required participants to be males and females between the ages of 18 and 65 who met at least 3 out of 4 criteria for PTSD according to the DSM-5, including hyperarousal symptoms. Exclusion criteria included recent or anticipated suicidal tendencies, specific psychiatric, neurological, or medical conditions, substance misuse, or having received recent treatment for PTSD in the last three months except when there was no trauma-related component and no change in treatment. Their PTSD symptom severity was assessed at three time points: pre-treatment, mid-treatment (after 10 sessions, end of day 5), and post-treatment, using the PTSD Checklist for DSM-5 (PCL-5). Furthermore, written informed consent was acquired from all participants in accordance with requirements from the Emory Institutional Review Board (STUDY0000038). This research was conducted at Emory University.

**Table 1.** Demographic and Clinical Characteristics.

| **Characteristics** |  | **Mean (SD) or N (%)** |
| --- | --- | --- |
| ***Demographic Characteristics*** |  |  |
| **Age** |  | 43.9(13.9) |
| **Sex** | **Female** | 17 |
|  | **Male** | 4 |
| **Race/ Ethnicity** | **White** | 16 |
|  | **Black** | 2 |
|  | **Mixed** | 1 |
|  | **Hispanic** | 1 |
|  | **Asian** | 1 |
| ***Clinical Characteristics*** |  |  |
| **PCL-5 Score** | **Pre-Treatment** | 37.0 (12.0) |
|  | **Mid-Treatment** | 33.4 (13.5) |
|  | **Post-Treatment** | 26.7 (9.9) |

### 2.2 EEG Data Collection

EEG data was collected from the individuals using EPOC X (Emotiv Inc, San Francisco, California), which is a 14-channel mobile EEG device. It provides EEG signal at AF3, F7, F3, FC5, T7, P7, 01, 02, P8, T8, FC6, F4, F8, and AF4. During the EEG data collection process, the EPOC X device’s wireless data transmission to real-time cloud storage compelled the choice of a 128 Hz sampling frequency, ensuring seamless integration within the study’s technological ecosystem ^21^. This choice ideally synchronizes the temporal dynamics of brain activity acquisition with the quick digital transfer of data to the cloud, ensuring no loss of essential information.

Each EEG sessions period, which consisted of two recordings, lasted approximately 3 minutes with participants’ eyes kept closed, yielded approximately 19,000 time points per sample. For each participant, EEG signals were recorded at three time points: pre-treatment, mid-treatment, and post-treatment, coinciding with the assessment periods for the PCL-5. In total, we acquired 38 EEG sessions from 20 participants, accounting for some sessions missing for certain individuals. To assess the reproducibility of the results, we conducted two EEG recording called “Recording A” and “Recording B” hereby (each approximately 180 seconds) for each participant at every time point.

### 2.3 PCL-5 Assessment

The assessment of PTSD symptoms was conducted using the PCL-5. This standardized self-report questionnaire consists of 20 items, each capturing the frequency and severity of specific PTSD symptoms over the past month. Participants were instructed to rate how much they have been bothered by each symptom in the past month (for pre-treatment) or the past week (for mid-treatment and post-treatment), employing a scale ranging from 0 (not at all) to 4 (extremely) ^22^.

### 2.4 Preprocessing

To ensure the quality and reliability of subsequent studies, the collected EEG data underwent extensive preprocessing implemented within MATLAB R2022b (The MathWorks Inc., Natick, MA, USA) harnessing the software’s computational capabilities and advanced toolsets for robust data analysis and interpretation. To successfully eliminate typical noise artifacts, the data were first re-referenced to the average reference. The raw data were then subjected to filtering operations to isolate relevant brainwave frequencies while attenuating noise. A low-pass Butterworth filter with a cutoff frequency of 55 Hz and a high-pass filter with a cutoff frequency of 0.01 Hz were used to reduce high-frequency noise and drift ^23^.

In refining the EEG data, wavelet independent component analysis (ICA) was employed to detect and separate independent sources contributing to the recorded EEG signals. To eliminate low-amplitude activities while retaining significant components, a wavelet thresholding approach was applied to the calculated independent components (ICs)^24^. This method involved dynamically determining the threshold value needed for effective wavelet thresholding, which was then subjected to scalar multiplication. This fine-tuned the reduction of background noise while preserving critical neural components.

### 2.5 EEG Variable Extraction

The primary focus of this study was to investigate the relationship between PTSD severity, quantified by PCL-5 values, and the complexity of EEG signals using Shannon entropy and spectral power across various electrophysiological bands.

Shannon entropy, a fundamental concept in information theory, is critical in assessing the complexity and unpredictability of EEG signals. It assesses the richness of information carried by brainwave amplitudes across different time points in the context of EEG analysis. Shannon entropy examines the distribution of amplitudes and converts them into probabilities by constructing the histogram of data from the EEG. These probabilities are then used to calculate the entropy value, which reveals the level of complexity in the EEG signal. This process is encapsulated by the equation

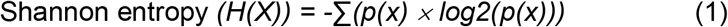

where, *H(X)* represents the Shannon entropy measure, *p(x)* represents the probability of observing the value *x* for Shannon entropy ^25^.

Spectral power variables are fundamental in analyzing EEG signals, providing insights into the distribution of signal power across different frequency bands. These variables quantify the amplitude information carried by brainwaves within specific frequency ranges, offering a glimpse into the neural dynamics underlying EEG data. In EEG analysis, spectral power is computed by segmenting the EEG signal into frequency bins and calculating the power within each bin. This is achieved through methods such as the Fast Fourier Transform (FFT) or similar techniques. The resulting spectral power values represent the strength of neural activity within distinct frequency bands, such as theta (4-8 Hz), alpha (9-12 Hz), beta (13-31 Hz) and slow-gamma (30-55 Hz) ^26^. These variables are pivotal for characterizing brain activity and can reveal patterns associated with cognitive processes and neurological conditions. The formula to compute spectral power (P) within a given frequency band is as follows:

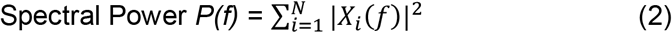

where, *P(f)* represents the spectral power in the frequency band *f, N* is the number of data points, *X*_*i*_*(f)* denotes the Fourier transform of the EEG signal at frequency *f* for the *i*^*th*^ data point. We used the Thomson multi-taper method from the Chronux toolbox (http://chronux.org/), implemented in MATLAB (Mathworks, Natick, MA) to calculate the spectral power in alpha band. The parameters for spectral analysis were the following: moving window= 5 s with 0.1 s overlap, the bandwidth produce TW=3, number of tapers K=5.

### 2.6 Statistical Analysis

To investigate the association between the extracted EEG variables and PCL-5 scores, we performed a partial correlation analysis with Pearson correlation, adjusting for age, sex, and batch as covariates. The batch variable was used to determine the association of samples with each participant. The p values for this analysis were corrected using a single false discovery rate (FDR) adjustment (p_FDR,4comparisons_<0.05) that included all regions of interest for each variable separately ^27^. To assess the reproducibility of the results, we conducted this analysis separately for “Recording A” and “Recording B” data. All data analysis and statistical computations were conducted using MATLAB (MathWorks, Natick, MA, USA) version R2022b.

## 3. Results

### 3.1 Shannon entropy from fronto-temporal regions link with PCL-5

Fig.1 illustrates the relationship between entropy and PCL-5 across four key EEG channels: AF3, AF4, T7, and T8 in “Recording A”. A negative association between Shannon entropy and PCL-5 was observed in AF3 (*r*= -0.456, FDR corrected *p*= 0.01), AF4 (*r*=-0.362, FDR corrected *p*= 0.04), and T7 (*r*= -0.472, FDR corrected *p*= 0.01), but not in T8. In the “Recording B”, similar trends in the correlation patterns between EEG channels and PCL-5 scores were observed primarily in AF3 (AF3, *r*= -0.168, uncorrected *p*=0.33), while the result was not significant. Additionally, a positive link between PCL-5 and Shannon entropy was noted in channels AF4 and T7, although none of these correlations were significant, even before FDR correction (Supplementary Fig.1).

**Figure 1.**
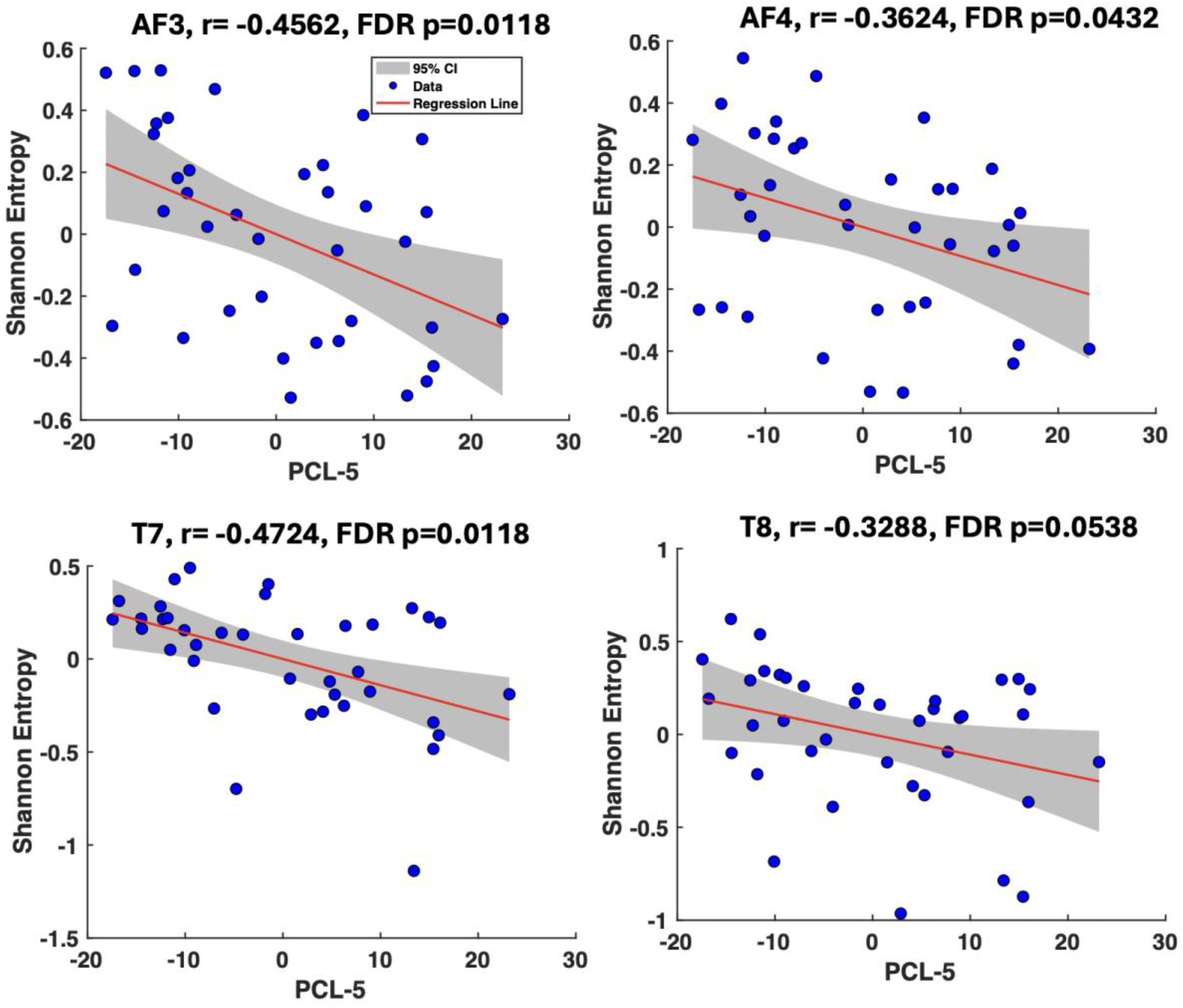
This figure displays the fronto-temporal EEG recordings’ Shannon entropy and its correlation with PCL-5 scores during “Recording A”, with each point representing an entropy-PCL-5 pairing adjusted for age, sex, and batch variables. The negative correlations at electrode sites AF3, AF4, and T7 are statistically significant, even after applying the False Discovery Rate (FDR) correction for multiple comparisons across these four regions of interest. This adjustment ensures that the likelihood of type I errors due to multiple testing is minimized. The persistent inverse relationships suggest that as EEG signal complexity increases, indicated by higher Shannon entropy, the severity of PTSD symptoms, as measured by PCL-5, tends to decrease.

### 3.2 Frontal alpha band power links with PCL-5

Fig. 2 illustrates the relationship between spectral power in the alpha band and PCL-5 scores across all four channels in “Recording A”. A significant negative correlation was observed between alpha band power and PCL-5 scores in AF4 (*r* = -0.429, FDR corrected *p* = 0.04), but not in other channels. Similarly, in “Recording B”, there was a negative correlation between the alpha band power from AF4 and PCL-5 (*r* = -0.369, uncorrected *p* = 0.02), but this association also did not reach significance after applying the FDR correction, as detailed in Supplementary Fig. 2.

**Figure 2.**
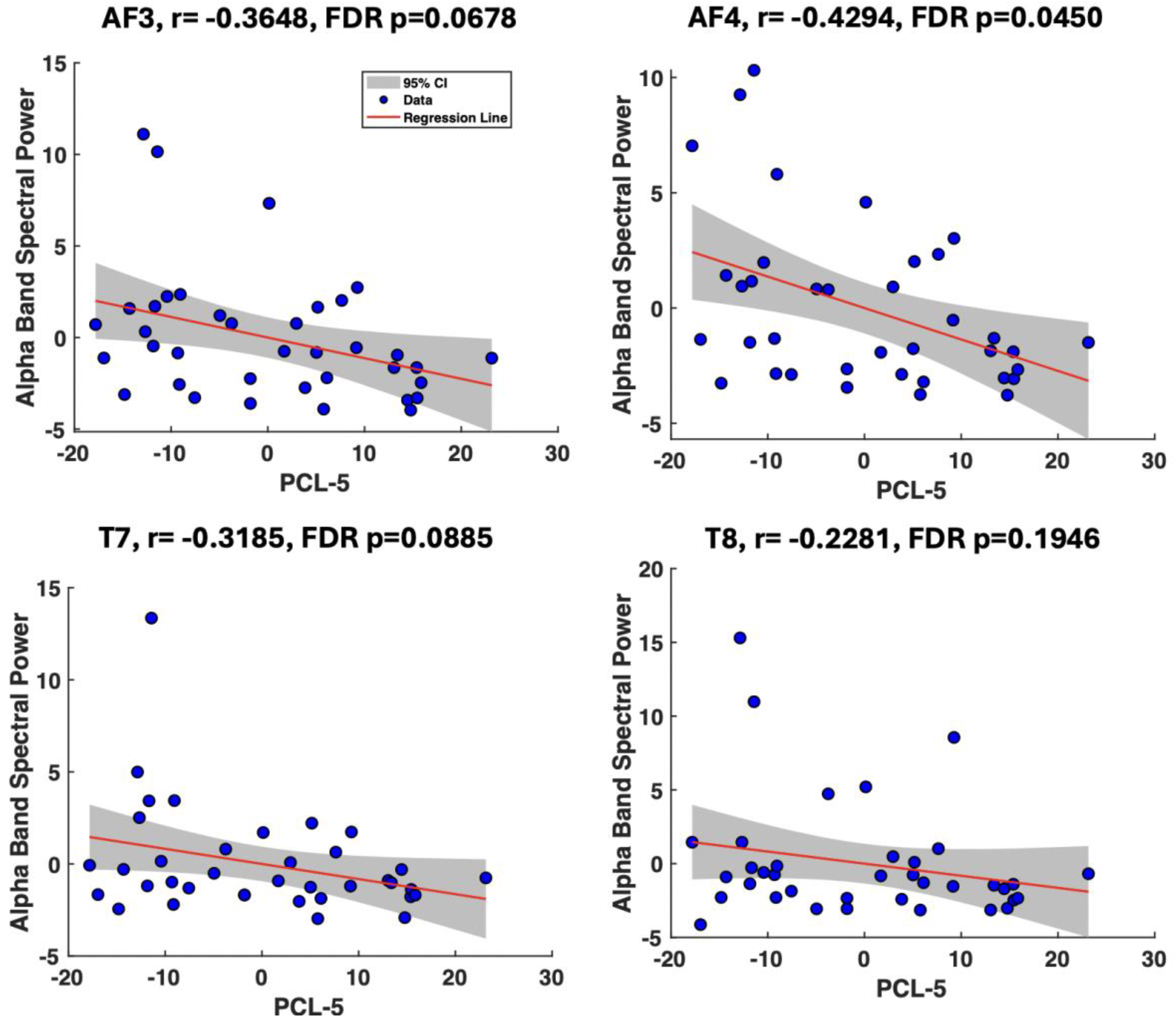
This figure displays the fronto-temporal EEG recordings’ alpha power and its correlation with PCL-5 scores during “Recording A”, with each point representing an alpha power-PCL-5 pairing adjusted for age, sex, and batch variables. The negative correlation at electrode sites AF3 is statistically significant, even after applying the False Discovery Rate (FDR) correction for multiple comparisons across these four regions of interest. This adjustment ensures that the likelihood of type I errors due to multiple testing is minimized.

## 4. Discussions

The current study assessed the capabilities of mobile EEG devices to explore the associations between entropy and alpha band power with PTSD symptom severity. While several studies have demonstrated an association between EEG signals and PTSD, here we highlight the potential of mobile EEG technology in associating EEG variables with PTSD symptom severity. Our EEG recordings from mobile devices revealed a negative link between fronto-temporal regions’ Shannon entropy and PTSD symptom severity. This indicates that individuals with greater PTSD symptoms exhibit reduced complexity and a more consistent pattern of brain activation in these regions, suggesting a decrease in the inherent irregularity typically associated with healthier EEG patterns. In other words, the reduced entropy in the fronto-temporal regions of PTSD sufferers reflects diminished neurophysiological variability, which may contribute to the symptomatic manifestations of PTSD, possibly including symptoms such as emotional and cognitive inflexibility. Our findings highlight Shannon entropy as a valuable metric for assessing the neurobiological impact of PTSD on brain function, offering the potential for both diagnostic advancements and targeted therapeutic interventions.

In our study, we also examined the spectral power within the alpha frequency band in relation to PTSD symptom severity over different timepoints. We observed that individuals with higher symptom severity exhibited lower alpha power, particularly in the frontal region. This negative association with the PCL-5 scores was consistent and reproducible across two recordings (i.e., Recording A and B). Previous studies have demonstrated reduced alpha power in individuals with PTSD using conventional EEG devices^19,28^. Our current findings align with these previous results, underscoring the reliability of mobile EEG devices in capturing similar patterns. Notably, the use of a mobile EEG allowed us to replicate these findings, thereby enhancing the practical applicability of EEG assessments in naturalistic settings and potentially broadening the scope for routine clinical and field use. Reduced alpha power is generally thought to represent decreased cortical inhibition and increased neural excitability. This consistency with earlier research not only validates our methodological approach but also contributes to the growing body of evidence supporting the role of alpha power reductions as a biomarker of PTSD symptom severity.

The fronto-temporal lobe, which plays a critical role in emotional processing, memory, and cognitive functions—all known to be compromised in PTSD—exhibited reduced brain complexity and alpha band power associated with higher PTSD symptom severity in our study^13^. In a study with 66 participants exposed to various mood inductions, amplified frontal EEG activity was linked with enhanced emotional regulation. This was evident by the reduced negative mood post-induction and decreased distraction from emotionally charged stimuli. Significantly, these findings were uniform across both brain hemispheres, emphasizing the multifaceted role of frontal EEG in mirroring emotional scenarios and regulatory capacities ^29^. Also, a study utilized EEG data from 40 students to specifically highlight the role of temporal regions, combining them with spectral and entropy biomarkers, resulting in comprehensive emotional profiles that shed light on the brain’s nuanced response to various emotional states^30^. Therefore, the observed correlations may signify a critical connection between altered neural dynamics in the fronto-temporal region and the expression of PTSD symptoms.

A recent study employing time-dependent Hurst analysis on EEG signals revealed that healthy controls (HC) exhibited greater complexity, as evidenced by the Hurst exponent in the F3 channel (frontal region). This significantly differentiated them from individuals with combat-related PTSD, highlighting the diagnostic potential of the Hurst exponent ^7^. In our research, we observed that participants with more severe PTSD symptoms displayed reduced EEG complexity; however, we utilized Shannon entropy as our measure rather than the Hurst exponent. We further investigated the relationship between the EEG Hurst exponent in the front-temporal regions in our cohort. Consistent with the prior study mentioned above, we observed that participants with higher PCL-5 scores exhibited lower Hurst exponents, indicating reduced complexity (see Supplementary Table1). While our results are not statistically significant, we found a consistent trend in both “Recording A” and “Recording B” that aligns with previous findings, particularly in the AF4 region. It is important to acknowledge the limitations due to our relatively small sample size and the differences in PTSD types (civilian vs. military). Future research with a larger sample size is required to validate the predictive power of the Hurst exponent with PCL-5 scores.

Mobile EEG devices have been utilized in previous studies for detecting neuropsychiatric disorders. For example, a recent study developed a model using single-channel, dry-electrode EEG technology to enhance the detection of depression intensity at specific moments^31^. Another study, utilizing EEG data from an Emotiv Epoc+ headset, developed a machine learning approach and attained an accuracy exceeding 98% in identifying depression in young adults through the analysis of distinct signal features ^32^. Another study introduced an efficient framework using resting-state EEG data and the MUSE EEG headband to accurately classify trait anxiety levels in participants, surpassing existing methods in precision ^33^. While our research, along with others, underscores the promising utility of mobile EEG devices in detecting neuropsychiatric disorders, more comprehensive future studies are crucial to evaluate and establish their diagnostic potential fully.

Our study’s limitations include a small sample, potentially limiting the generalizability of findings. The sex imbalance, with a higher representation of females, introduces a sex-related bias, and future studies should strive for a more balanced distribution. The predominantly White American composition of our sample may restrict the applicability of results to diverse populations. A more comprehensive understanding of mobile EEG-derived biomarkers and PTSD symptoms would benefit from larger, more diverse samples that include participants from various ethnic backgrounds and consider sex-related differences. Moreover, the lower sampling frequency of our study restricts our ability to assess the utility of higher frequency band spectral power in relation to PCL-5 scores. Finally, the study is part of an ongoing clinical trial in which 50% of participants receive active TMS and 50% sham. For this reason, we only explored the association with symptoms here, but further evaluation of the effect of treatment will be performed when the clinical trial is completed.

In conclusion, this research advances the field of EEG analysis by emphasizing the importance of the fronto-temporal lobe, unravelling the complexities of EEG temporal dynamics, and highlighting the potential of mobile EEG to predict PTSD. While our findings may offer some significant insights, they represent an initial step towards a deeper understanding of the neural mechanisms underlying the clinical phenomena studied and provide proof-of-concept for these approaches. Further studies, particularly with expanded sample sizes, are essential to more clearly define the complex relationships in EEG data and their relevance to our research goals.

## Data Availability

All data produced in the present study are available upon reasonable request to the authors.

## 5. Acknowledgements

We express our gratitude to the individuals responsible for data collection, whose meticulously efforts were critical to the study’s success. We also appreciate the participants in the research for their substantial contributions, as well as our funding sources for their assistance.

## 8. Author’s Contribution

AP developed the study, conducted data analysis, interpreted the results, and wrote the original manuscript draft. MS developed and supervised the study, interpreted the results, and edited the original manuscript draft, provided critical review to the initial draft, and secured funding. SM collected the data and provided critical review to the initial draft. CH collected the data and provided critical review to the initial draft. EW assisted in data cleanup and analyses and provided critical review to the initial draft. KJR provided critical review to the initial draft. VC provided critical review to the initial draft and secured funding. SVR developed and supervised the study, collected the data, interpreted the results, and edited the original manuscript draft, provided critical review to the initial draft, and secured funding for the overall clinical trial.

