## Supplementary Material for "Towards predicting PTSD symptom severity using portable EEG-derived biomarkers"

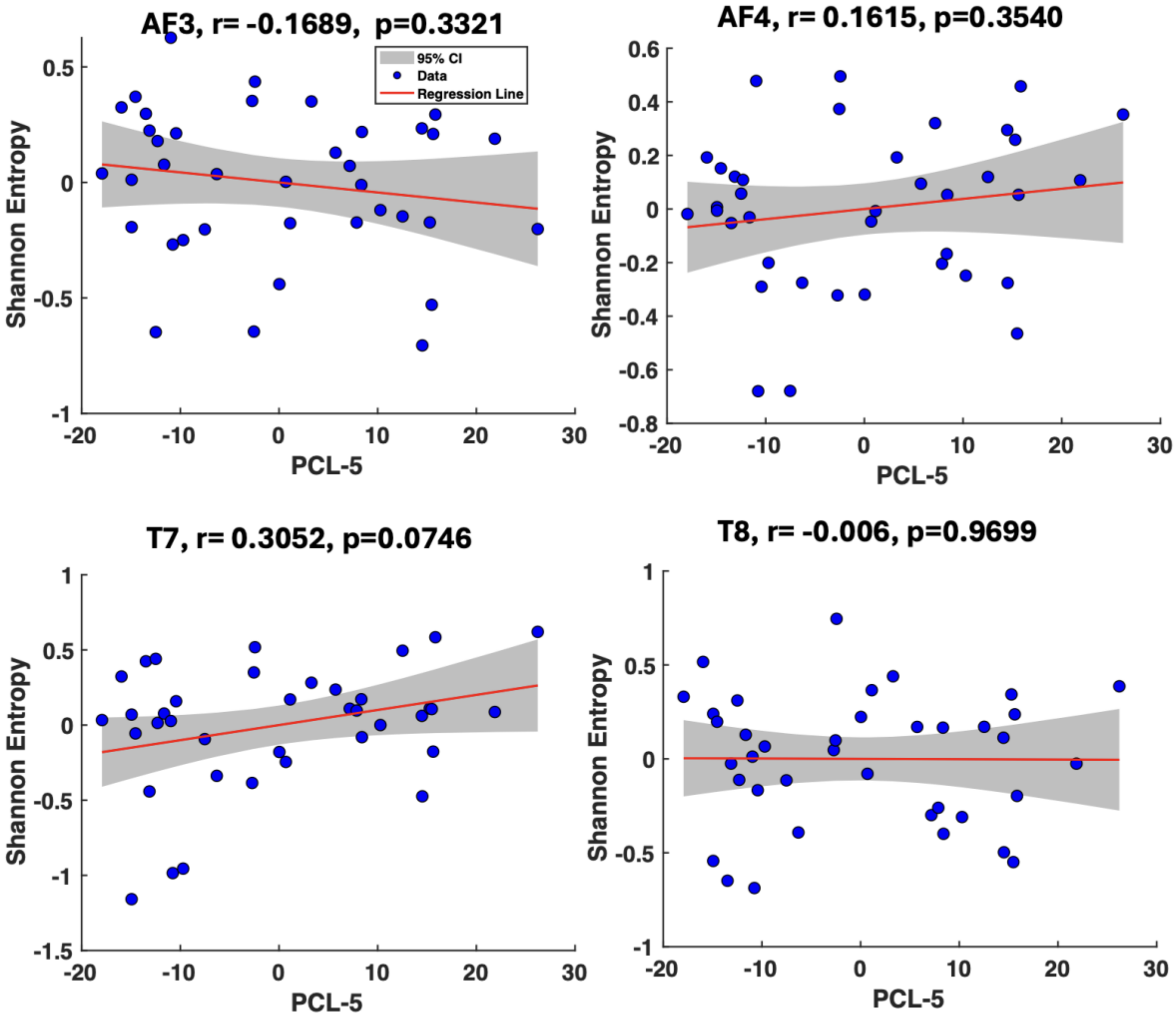


**Supplementary Figure. 1.** This figure illustrates the Shannon entropy from fronto-temporal EEG recordings and its correlation with PCL-5 scores during "Session B". Each data point represents an adjusted pairing of entropy and PCL-5 scores, accounting for age, sex, and batch variables. Despite the observed negative correlation between Shannon entropy and PCL-5 scores in the AF3 region during Session A, this link was not significant in "Session B".

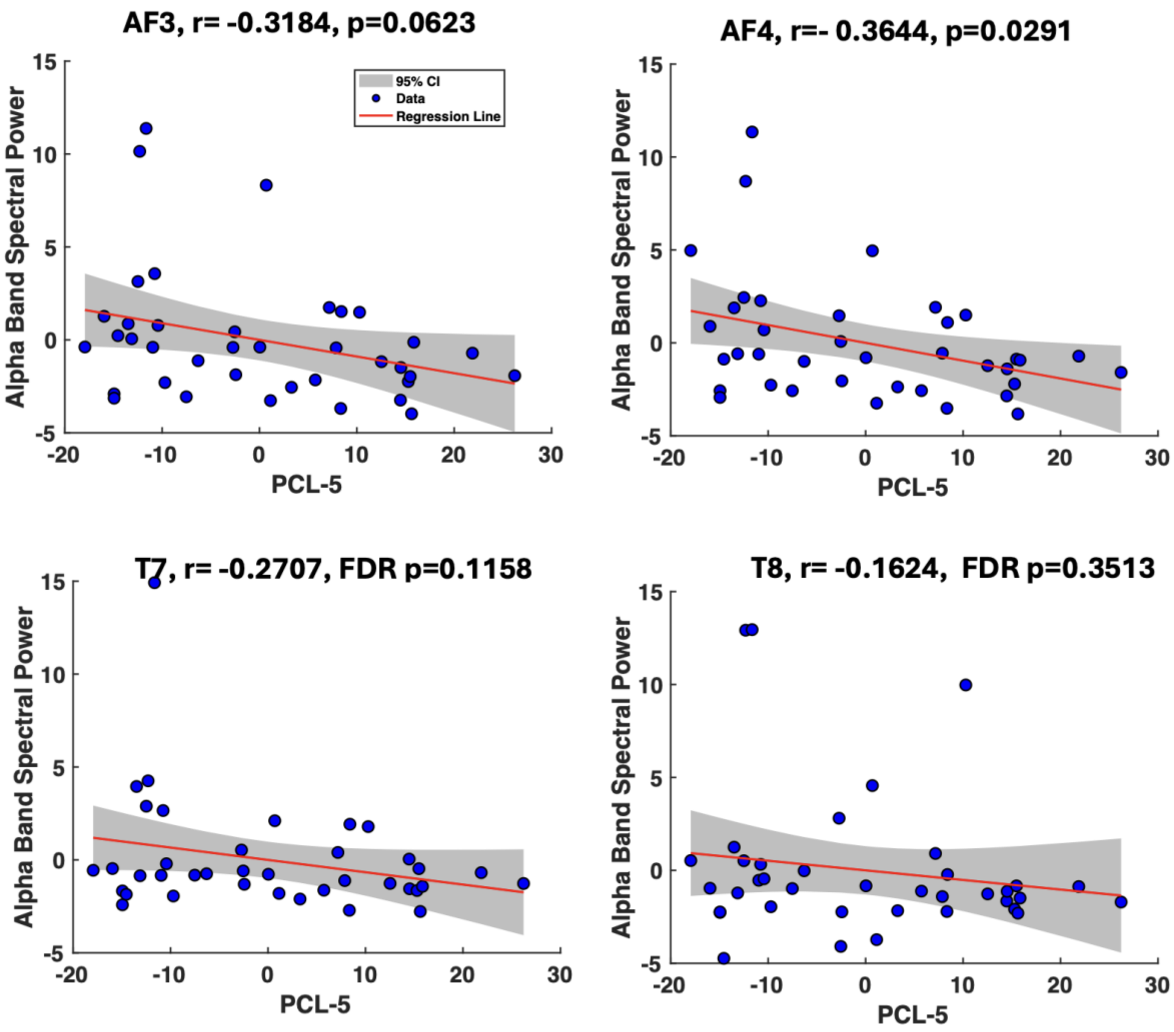


**Supplementary Figure. 2.** This figure presents the alpha band power from fronto-temporal EEG recordings and its correlation with PCL-5 scores during "Session B". Each data point represents an adjusted pairing of alpha band power and PCL-5 scores, accounting for age, sex, and batch variables. A significant correlation between alpha band power and PCL-5 scores was observed in the EEG AF4 region during "Session B", similar to "Session A" (uncorrected $p < 0.05$). However, this correlation was not significant in Session B after applying FDR correction.

**Hurst Exponents link with PCL-5:** In our study, we harness the power of Hurst exponents to probe the temporal complexity embedded within EEG signals. Similar to Shannon entropy's role in assessing information richness, the Hurst exponent quantifies long-range correlations and self-similarity in time series data. By analyzing EEG data across various channels and segment lengths, we unveil the temporal intricacies that may underlie our research objectives. The Hurst exponent offers a unique perspective on the fractal nature of neural oscillations, providing valuable insights into the temporal dynamics of our EEG recordings. This analysis enriches our understanding of the neural processes associated with the clinical phenomena under investigation.

The formula for estimating the Hurst exponent is expressed as follows:

$$\text{Hurst Exponent } (H) = \lim (T\to\infty)\ [\log(R/S(T))] / \log(T) \qquad (1)$$

**Supplementary Table 1: Channel-Wise Correlation Analysis of Hurst Exponents with PCL-5 for "Session A" and "Session B"**

| Channels | R-value (uncorrected p-value) | |
|---|---|---|
| | **Session A** | **Session B** |
| **AF3** | 0.004 (0.981) | -0.248 (0.152) |
| **T7** | 0.106 (0.540) | -0.198 (0.255) |
| **T8** | -0.060 (0.727) | -0.213 (0.220) |
| **AF4** | -0.070 (0.684) | -0.086 (0.624) |
